# Persistence of geographic barriers to maternal care services following a health system strengthening initiative in rural Madagascar

**DOI:** 10.1101/2023.10.20.23297294

**Authors:** Felana A. Ihantamalala, Mauricianot Randriamihaja, Ann C. Miller, Rado J.L. Rakotonanahary, Mbolatiana Raza-Fanomezanjanahary, Jacques Aubin Kotchofa, Marius Randriamanambintsoa, Haja Ramarson, Benedicte Razafinjato, Vololoniaina Rasoanandrasana, Matthew Bonds, Karen E. Finnegan, Andres Garchitorena

## Abstract

**Background:** Geographic access to healthcare continues to pose a significant challenge for pregnant women in rural areas of sub-Saharan Africa, resulting in consistently high rates of maternal mortality. Geographic barriers can persist even in settings where financial barriers have been reduced and health system strengthening (HSS) efforts are in place. The aim of this study is to combine analyses of a population-representative cohort and geolocated maternal consultation data at the village level to gain a precise understanding of spatiotemporal changes in the utilization of maternal care services in a rural district of Madagascar benefiting from HSS support.

**Methods:** We collected monthly information on antenatal care visits, deliveries and postnatal visits from the registries of 18 primary care health centers in Ifanadiana District, from 2016 to 2018. Similar data were collected from a district-representative cohort via surveys on over 1500 households done in 2016 and 2018. We estimated precise travel time from each village to the nearest health center to understand spatio-temporal variations in maternal care access, and to assess the impact of geographic barriers via statistical analyses.

**Results:** Women who lived within a one-hour walk from a health facility in the HSS catchment area had rates of per capita utilization of most maternal health services were roughly twice that those who lived 1-2 hours away and three times higher than those who lived over 2 hours away. The exception was the first antenatal care visit (ANC1), for which travel time had more modest effect. Improvements to primary care services due to HSS in this setting were only observed among women living within two hours from health centers. Statistical models revealed that women’s travel time from a health facility was the strongest determinant of maternal care service utilization.

**Conclusion:** This study shows how a combination of geo-located health system information and population-representative data can help assess the impact of geographical barriers to maternal care in rural areas of sub-Saharan Africa. It highlights that women who live more than 2 hours from a health facility had virtually no access to maternal health services despite efforts in place to reduce financial barriers to care and strengthen the health system.

## Background

There have been significant improvements in maternal care across the world in recent decades, with increased access to antenatal care (ANC), skilled birth attendance, and postnatal care. Maternal mortality rates have decreased by nearly 40% globally between 2000 and 2017 [1]. Despite this, inadequate access to quality maternal care services remains a critical issue, especially in the WHO Africa region [2,3]. Financial and geographical barriers, including long travel distances to health facilities, transportation limitations, and poor infrastructure, can pose formidable obstacles for pregnant women seeking essential care. This can lead to inadequate monitoring of pregnancies, missed opportunities for early intervention, and increased risks for complications during childbirth [2,4,5]. In addition, social and cultural factors, such as gender inequality, traditional practices, and societal norms, can influence maternal care-seeking behaviors and contribute to disparities in care [6,7]. Addressing these multifaceted challenges requires comprehensive strategies that encompass strengthening health systems, improving access to services, empowering women, and promoting equitable and culturally sensitive care for all expectant mothers.

Geographic barriers to health facilities are particularly persistent in rural areas of sub-Saharan Africa, where health system infrastructure is sparse and pregnant women have to travel many hours, often on foot, to reach the nearest facility [8–10]. Nearly one third of women of child bearing age are located more than 2-h travel time from the nearest hospital [11]. A review of studies on maternal health access conducted in several Sub-Saharan countries revealed that geographical barriers were the most common factor reducing the use of maternal care services [2,11]. For example, in Zambia, each 10 km increase in distance from a health center decreased the likelihood of receiving antenatal care by 25% [12]. Long distances can also prevent pregnant women from giving birth assisted by health professionals and therefore increase the risk of death from medical complications, such as sepsis and severe venal hemorrhaging, that result from childbirth [13,14]. In a study in Nigeria, distance was found to be the second main factor for not receiving postnatal care [15]. Geographic barriers to maternal care can remain even in settings where financial barriers have been reduced and the health system has been strengthened [16,17], which poses challenges for reaching universal health coverage.

Geographic barriers to health care are especially acute in Madagascar [18], where 60% of women make complete 4 antenatal care visits, and less than 40% of women deliver at a health facility, with even lower access in rural areas of the country [19]. This is associated with one of the lowest health expenditures per capita in the world (18 USD in 2020), and a limited number of health professionals, at less than two doctors per 1,000 people [20]. In Ifanadiana where this study was conducted, nearly 70% of the population live more than 5km from a primary care facility. Utilization of outpatient consultations were found to be six times greater for those who live near primary care center versus those who live beyond 5km.

In 2014, the Ministry of Public Health (MoPH) established a roadmap to reduce maternal mortality to 300 per 100,000 live births by 2019 by ensuring the provision of essential, integrated, quality services around pregnancy and childbirth, with an emphasis on adolescent and youth health [21]. Yet, maternal mortality has remained high and relatively stable over the past two decades at levels over 400 per 100,000 live births [22,23]. While the impact of socio-cultural perceptions, financial barriers and health system factors on maternal care use have been previously described in Madagascar [4,24,25], there is no in-depth evaluation of the role of geographic barriers. This evaluation could help the MoPH understand the extent of the problem and devise appropriate strategies to overcome them.

In this study we assessed whether geographic access to maternal care services remains a major barrier following a health system strengthening initiative in a rural health district of Madagascar. The initiative, implemented since 2014 in partnership between the Ministry of Public Health and the NGO Pivot aims to create a model system of a strengthened public health system at all levels of care (community, primary healthcare, and hospital) through the integration of health system readiness support (infrastructure, human resources, materials), financial coverage, and vertical improvement of clinical programs. In its first 4 years, the intervention was associated with a relative increase of 20-30% in antenatal, delivery and postnatal care, but absolute levels remained low [26]. Here, we combine analyses of a population-representative cohort and geolocated maternal consultation data at the village level to gain a precise understanding of spatiotemporal changes in the utilization of maternal care services in Ifanadiana district 2 years after the implementation of the intervention, with a focus on geographic barriers.

## Methods

### Study area

The study was conducted in Ifanadiana, a rural district located in southeastern Madagascar, in the region of Vatovavy. The district covers an area of 3975 km² and has a population of about 200,000. During the study period, the district’s health system had one district hospital and 19 public health centers that provide primary health care services to the local population, including 1 major health center (CSB2) in each of the district’s 13 communes, and 6 more basic health centers (CSB1) in communes that are too large to be covered by only one health center. The landscape is characterized by rolling hills, dense forests, and a multiplicity of rivers and streams that flow through the district. There is only one paved road crossing the district from West to East and two non-paved axes connecting the main towns in the North and South of the District, which are partly accessible by 4WD vehicles or all-terrain motorcycles. Most villages in the district are connected to each other by small paths only accessible by foot. The lack of accessible roads and reliable means of transportation limit the ability of pregnant women to access maternal care services.

In 2014, only one-third of women completed the recommended 4 antenatal care visits, less than one in five women gave birth at a health facility, and the mortality rate was more than twice the national average, at 1044 deaths per 100,000 live births [27]. To address these challenges, the HSS initiative implemented by the MoPH and Pivot aims to expand healthcare infrastructure, increase the number of healthcare professionals, and strengthen healthcare programs. The initiative was initially rolled out in 4 communes (administrative unit that typically ranges from 6,000 to 30,000), and expanded to one additional commune in 2017. Activities on maternal care over the first 4 years of implementation (2014-2018) focused on building the health system capacity to provide high-quality, comprehensive care. This included training of health care workers in emergency obstetric and neonatal care, provision of essential medical equipment and supplies to health facilities, removal of user fees at all levels of care, and community-based interventions to increase sensitization and awareness around maternal care and family planning. This program is currently expanding to all communes of the district, with an agreement between the government of Madagascar a Pivot to expand across two more districts (with a total of one million people), starting in 2024.

### Data collection

#### Health system data

For this study, data was gathered on maternal care consultations from January 2016 to December 2018 for 18 out of 19 health centers in Ifanadiana District. To obtain the geographic residence of each woman attending health centers for a maternal care visit, information was gathered at the individual level from prenatal care and delivery registers.

The antenatal care register includes the age of each pregnant woman, their village of residence, and the number of her antenatal care visit (1 to 4 visits), and the delivery register includes the age of each woman who gives birth at the facility, their village of residence, as well as information on postnatal care visits. To create a digital database, pictures were taken from both registries at each health facility during field missions between July and August 2020, and the information in these pictures was entered manually into an electronic database. One health center had misplaced its maternal care registers for that period and these data could not be retrieved.

Population estimates for each fokontany (smallest administrative unit comprising a village or small group of villages) were obtained from the MoPH. In addition, the number of medical staff at each primary care facility (PHC) over time within the study period was obtained from internal data from the NGO Pivot, and included the number of nurses and doctors at each PHC per year. The use of health system data was authorized by the MoPH.

#### Population survey data

Population-level data on maternal care was obtained for 2016 and 2018 from the Health Outcomes and Prosperity longitudinal Evaluation (IHOPE), a longitudinal cohort study initiated in Ifanadiana district in 2014. The study involves a series of cross-sectional surveys conducted in a representative sample of 1600 households, which are revisited every 2 years. The questionnaires used in the surveys are based on the internationally validated Demographic and Health Surveys (DHS). The sample is stratified in two groups: the initial Health System Strengthening (HSS) catchment and the Rest of the District (RoD). A total of eighty clusters, half from each stratum, were randomly selected from enumeration areas mapped during the 2009 census. Within each cluster, twenty households were randomly selected for participation in the survey. Data collection and survey coordination are carried out by the Madagascar National Institute of Statistics (INSTAT). During the surveys of 2016 and 2018, 1514 and 1512 households participated, respectively.

The survey included a household questionnaire and individual questionnaires for men and women of reproductive age (15-59 years for men; 15-49 years for women). All eligible women and men residing in the sampled households, whether permanent residents or visitors, were interviewed. The information gathered in the questionnaires included indicators of socioeconomic status (education, employment, household durable assets); indicators of current illness (for the last 30 days); preventive behaviors (bed net ownership, access to water and sanitation); women’s reproductive histories and care-seeking behaviors for reproductive health; and children’s health, development, and care-seeking.

#### Geographic data

We used data on distance and travel time from each household in Ifanadiana district to the nearest Primary Health Center (PHC) produced in a previous study [28]. A detailed description of the methods can be found in Ihantamalala et al. (2020). In brief, a full mapping of the district was conducted in 2018 via OpenStreetMap (OSM), including all footpaths, residential areas, buildings and rice fields. For this, the district was divided into 1km² tiles and mapped using very high-resolution satellite images. The mapping was done in two steps: one user performed the mapping of all geographic entities in a tile, and subsequently, another user validated the mapping to ensure the quality of the data. This resulted in over 100,000 buildings and 20,000 km of footpaths mapped. The Open-Source Routing Machine (OSRM) engine was then used to accurately estimate the shortest path and distance between each house in the district and the nearest PHC. In parallel, to obtain accurate estimates of travel time, we conducted field tracking of 168-foot itineraries covering nearly 1000 km in Ifanadiana, recorded via the OsmAnd App. This information was combined with remotely sensed land cover, climate and elevation data to statistically model travel speed associated with different terrain characteristics and climatic conditions via a generalized additive mixed model. Finally, using this model in combination with the shortest paths obtained from OSRM, we predicted travel time to reach the nearest Primary Health Center (PHC) for each household in Ifanadiana. Travel time for each fokontany (health system data) or for each survey cluster (cohort data) represents the average travel time of all the buildings in that area.

### Data analyses

#### Health system analyses

Patient-level data on ANC1, ANC4, deliveries and postnatal care from each health facility was aggregated by fokontany and month. To obtain an estimate of “per capita” utilization, the total number of maternal care consultations was divided by the corresponding population of expected pregnant women (for ANC1 and ANC4) or expected births (for deliveries and postnatal care) in the fokontany. In line with MoPH estimates, the expected number of pregnant women and the expected number of deliveries were set at 4.5% and 4% of the total population of each fokontany, respectively.

Spatial and temporal variations in each maternal care indicator were assessed from 2016 to 2018. The linear and non-linear impact of travel time from each fokontany to the nearest health center on maternal care utilization rates was investigated, to understand how geographic barriers impact the utilization of health services by pregnant women in the district. The monthly and annual fluctuations in utilization rates for these maternal care indicators were also assessed across different travel time categories: less than one hour, between one and two hours, and over two hours. Finally, a time-series analysis was carried out on the monthly time series to characterize the relative change in consultations by travel time category while controlling for health system factors and time trends. For this, a negative binomial regression was used in a generalized linear mixed model framework, with the fokontany of the patient set as a random intercept. Health system factors included the number of health staff per facility over time, the type of PHC (CSB1 vs CSB2) and whether the PHC was supported by the HSS initiative. Time trends included a linear annual trend and a lagged consultation trend (1-month). A separate model was done for each maternal care indicator. All analyses were conducted with R software, version 4.1.3 and R package ‘lme4’ [29]. Maps were done with ArcGIS, version 10.8.

#### Population survey analyses

Equivalent analyses to those described for health system data were carried out for survey data. For the estimation of maternal care indicators at the population level for each survey year, 2016 and 2018, the last birth of every participating woman that occurred within the previous two years was included. The spatial distribution of maternal care access was obtained by estimating the average in each of the 80 spatial clusters for both surveys. The linear and non-linear impact of travel time on maternal care access, and changes over time by travel time category were also assessed. Similar to statistical analyses of health system data, a logistic regression model was used to characterize the odds ratio in maternal care access associated with travel each time category while controlling for health system factors (number of health staff per facility over time, the type of PHC, and whether the PHC was supported by the HSS initiative) and time trends (linear annual trend). Sampling weights that adjusted for unequal probability of selection due to stratification and non-response were calculated for household, women’s and men’s surveys. Estimates were done using survey commands available in R-package *survey* and applicable sampling weights [30].

## Results

### Spatial and geographic trends in maternal care access

Between 2016 and 2018, 25,555 women attended a PHC for an ANC1, 7,050 for an ANC4, 7,553 for delivery and 4678 for postnatal care in the 18 health centers of Ifanadiana District (Table 1). Of 244 women surveyed who had a birth in the previous two years, 78.11% reported coming to the health center for ANC1, 46.02% for ANC4, 27.70% for delivery and 26.33% for postnatal care. Spatial analyses revealed that ANC1 utilization was substantially higher and more evenly distributed than other maternal care indicators, with 43.55% of fokontany having over 0.75 per capita visits per year (Figure 1A). Low rates of ANC1 were only observed in some remote areas of the northeast and northwest of the district. In contrast, utilization for ANC4, delivery and postnatal care was almost exclusively concentrated in close proximity to health centers, and fewer than 9.6% of fokontany had per capita utilization over 0.5 per year (Figure 1A, Additional file1). Spatial patterns observed from health system data were consistent with those from cohort surveys for all indicators (Figure 1B). For instance, while 65% of survey clusters had at least 75% of pregnant women surveyed reporting having attended a health center for ANC1, less than 36% of clusters had levels of health care access over 50% for any of the other indicators, all of them located in close proximity to health centers (Figure 1B, Additional file2).

**Figure 1:**
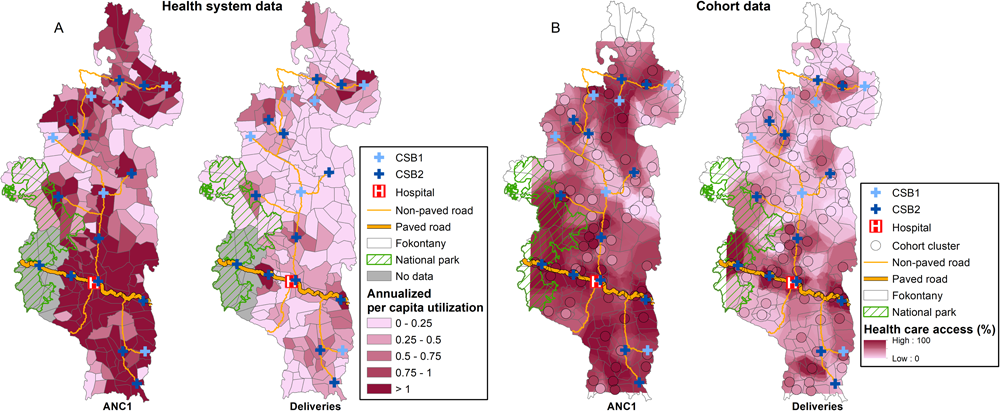
Spatial variation in antenatal care (ANC1) and deliveries in Ifanadiana District, 2016-2018. A) Annualized per capita utilization out of expected pregnant women, estimated as the fokontany average from data of all health centers. B) Percentage of pregnant women included in each of 80 clusters in cohort surveys (black circle) who accessed each maternal care service, with values interpolated using inverse distance weighted interpolation.

**Table 1:**
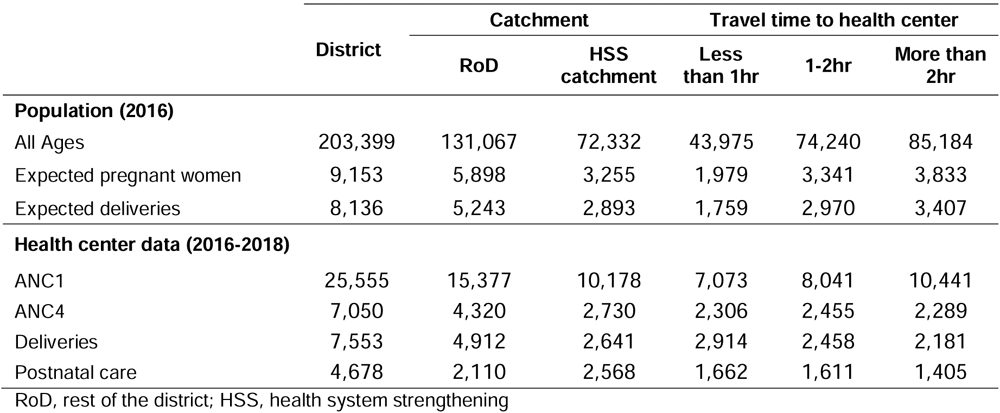
Description of population size and use of maternal care services, from health system data.

Geographic trends in maternal care access were observed for all indicators, with slight differences between the HSS intervention catchment and RoD (Figure 2). There was a consistent decline in utilization rates for women living at increasing travel time from health centers, but women in the HSS catchment who lived in close proximity to health centers had higher rates of maternal care access than those in the RoD. The decrease was particularly pronounced for ANC4, deliveries and postnatal care, were we observed an exponential decline as a function of travel time. Annual per capita utilization rates for women living over 2 hours from a PHC was consistently under 0.25 for ANC4, deliveries and postnatal care in both catchment areas (Figure 2A). Maternal care access from cohort data revealed similar patterns, with a smaller decline for ANC1 than for the rest of maternal indicators (Figure 2B). The decline in women’s access to PHC was particularly pronounced within the first 2 hours of travel time from the facility. Beyond this point, ANC4 access remains stable at about 25% for women in the HSS catchment, while it continues to decrease for deliveries and postnatal care until access is close to zero for women living further than 3 hours from a PHC (Figure 2B). For women in the RoD, lower access overall meant that geographic trends were less pronounced.

**Figure 2:**
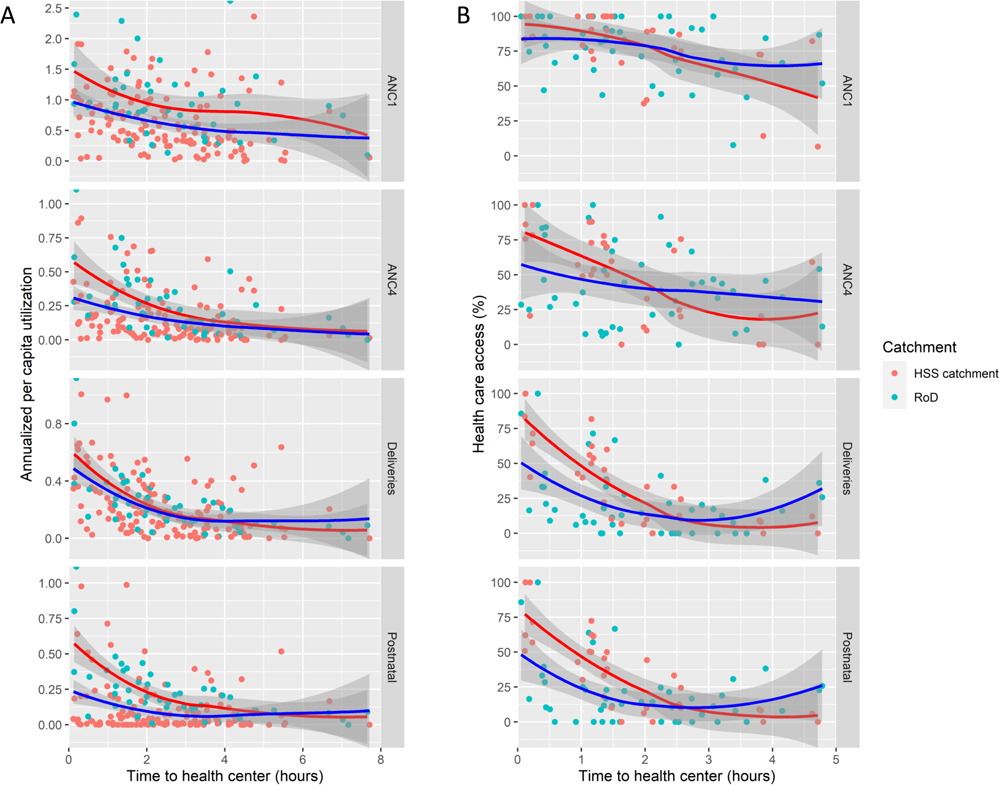
Impact of travel time to health centers on maternal care access in Ifanadiana District, 2016-2018. A) Annualized per capita utilization out of expected pregnant women, for antenatal care (ANC1, ANC4), deliveries and postnatal care, estimated as the fokontany average (points) from data of all health centers. Solid lines represent the non-linear smooth of the relationship between utilization and travel time, inside the HSS catchment (red) or in the rest of the district (RoD, blue) B) Percentage of pregnant women included in each of 80 clusters in cohort surveys (points) who accessed each maternal care service. Solid lines represent the non-linear smooth of the relationship between access and travel time, inside the HSS catchment (red) or in the rest of the district (RoD, blue)

#### Impact of geographical barriers to maternal care access

Temporal trends as a function of travel time to health facilities were relatively stable for all four indicators of maternal care access during the study period (Figure 3). In the HSS catchment, women residing in fokontany within an hour distance had rates of per capita utilization roughly twice that of those who lived 1-2 hours away and three times higher than those who lived more than 2 hours away (Figure 3A). The only exception was ANC1, where these differences were smaller. Women living over two hours away had the lowest utilization rates, at under 0.2 per capita-year for ANC4, deliveries and postnatal care, with similar trends in both the HSS intervention catchment and the RoD. We observed similar patterns in cohort data, where women residing within 1 hour had substantially higher levels of health care access in the HSS catchment, consistently over 75% for ANC1 and ANC4, and over 50% for deliveries and postnatal care for both survey years (Figure 3B). Consistent with analyses of health system data, women living over 2 hours from a PHC had levels of health care access under 25% for ANC4, deliveries and postnatal care, regardless of HSS support. However, for those from the RoD, access for women who lived 1-2 hours from a PHC was relatively similar to those who lived over 2 hours (Figure 3B).

**Figure 3:**
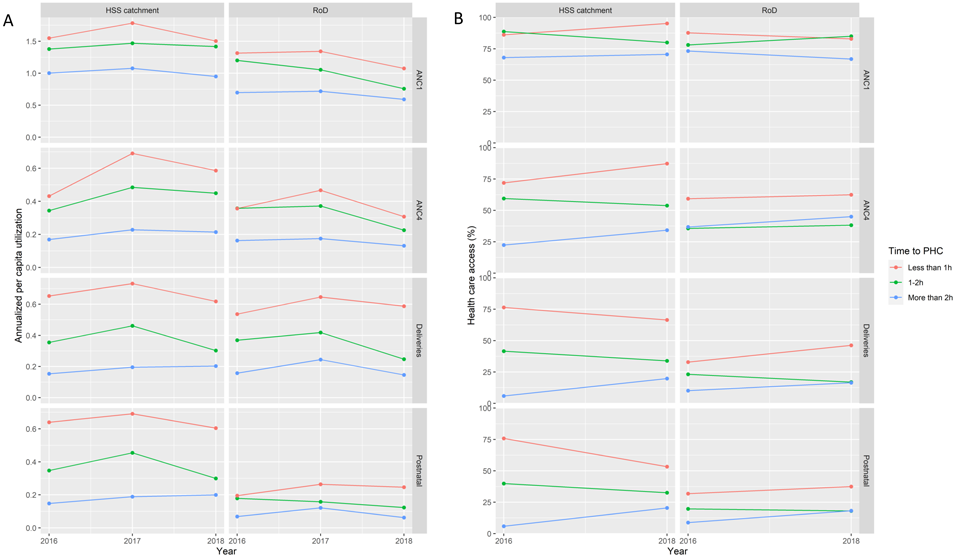
Annual variation in maternal care access by distance to health center in Ifanadiana District, 2016-2018. A) Annualized per capita utilization out of expected pregnant women, for antenatal care (ANC1, ANC4), deliveries and postnatal care, estimated as the average of all fokontany in 3 categories of geographic access (Less than 1 hour to PHC, 1-2 hours, more than 2 hours) B) Percentage of pregnant women included in cohort surveys who accessed each maternal care service, grouped in 3 categories of geographic access (Less than 1 hour to PHC, 1-2 hours, more than 2 hours)

After accounting for health system factors, temporal trends and HSS support, our statistical models revealed that women’s travel time from a PHC was the strongest determinant of maternal care service utilization for all four indicators (Table 2 and 3). The number of staff, the type of health center, and HSS support were not significantly associated with women’s per capita utilization of healthcare centers overall, with the only exception of ANC4 (Table 2). Living 1-2 hours from a PHC was associated with a relative decrease ranging from 0.62 for deliveries to 0.86 for ANC1, as compared with those living within 1 hour. For those living further than 2 hours, the relative change ranged from 0.4 for deliveries to 0.72 for ANC1. The impact of geography was also evident in statistical analyses of cohort data (Table 3), but associations were only statistically significant for women living more than 2 hours away from a PHC. Women living further than 2 hours, had an odds ratio of health care access ranging from 0.13 for deliveries to 0.3 for ANC, as compared with those living within 1 hour. In these models, the number of health staff working at the PHC was associated with a positive impact on deliveries and postnatal care.

**Table 2:** Multivariate analyses of health system data (generalized linear mixed model, negative binomial distribution)

| Variables | ANC1<br>Relative change<br>(95% CI) | ANC4<br>Relative change<br>(95% CI) | Deliveries<br>Relative change<br>(95% CI) | Postnatal<br>Relative change<br>(95% CI) |
| --- | --- | --- | --- | --- |
| **Geographic factors** |  |  |  |  |
| Travel time (*ref. less than 1h*) |  |  |  |  |
| 1-2 hr | 0.84 (0.78-0.90) *** | 0.78 (0.68-0.89) *** | 0.60 (0.53-0.67) *** | 0.65 (0.56-0.76) *** |
| More than 2 hr | 0.72 (0.67- 0.77) *** | 0.53 (0.46- 0.60) *** | 0.40 (0.36-0.45) *** | 0.47(0.40-0.54) *** |
| **Health system factors** |  |  |  |  |
| Number of health staff | 1.03 (0.91-1.18) | 0.89 (0.69-1.13) | 1.01 (0.84-1.20) | 0.97 (0.73-1.27) |
| PHC level (*CSB2 vs. CSB1*) | 1.41 (0.95-2.09) | 2.52 (1.24-5.11) * | 1.18 (0.78-1.79) | 1.41 (0.57-3.45) |
| HSS catchment (*RoD vs. HSS catchment*) | 0.97 (0.83-1.14) | 1.18 (0.84-1.67) | 0.99 (0.72-1.35) | 0.74 (0.51-1.09) |
| **Temporal trends** |  |  |  |  |
| Annual linear trend | 0.94 (0.91-0.97) *** | 1.00 (0.94-1.06) | 0.93 (0.88-0.99) * | 0.96 (0.89-1.03) |
| Lag (1-month) | 1.35 (1.33-1.38) *** | 1.93 (1.81-2.07) *** | 1.57 (1.49-1.65) *** | 2.01 (1.86-2.18) *** |

\*p <0.05; \*\*\*p <0.001
PHC, primary health center; HSS, health system strengthening; RoD, rest of the district.

**Table 3:** Multivariate analyses of cohort data (generalized linear model with applicable survey weights, binomial distribution)

| Variables | ANC1<br>Odds Ratio<br>(95% CI) | ANC4<br>Odds Ratio<br>(95% CI) | Deliveries<br>Odds Ratio<br>(95% CI) | Postnatal<br>Odds Ratio<br>(95% CI) |
| --- | --- | --- | --- | --- |
| **Geographic factors** |  |  |  |  |
| Travel time (*ref. less than 1h*) |  |  |  |  |
| 1- 2 hr | 0.87 (0.36-2.12) | 0.45 (0.2-1.02) | 0.46 (0.21-1.04) | 0.52 (0.23-1.19) |
| More than 2 hr | 0.3 (0.12-0.73) * | 0.27 (0.13-0.56) *** | 0.13 (0.06-0.26) *** | 0.17 (0.08-0.37) *** |
| **Health system factors** |  |  |  |  |
| Number of health staff | 1.36 (0.86-2.15) | 1.3 (0.93-1.81) | 1.51 (1.11-2.06) * | 1.5 (1.09-2.06) * |
| PHC level (*CSB2 vs. CSB1*) | 2.36 (0.9-6.19) | 1.78 (0.73-4.36) | 2.14 (0.88-5.24) | 1.83 (0.68-4.93) |
| HSS catchment (*RoD vs. HSS catchment*) | 1.29 (0.51-3.27) | 1.37 (0.59-3.18) | 1.32 (0.55-3.13) | 1.26 (0.53-2.97) |
| **Temporal trends** |  |  |  |  |
| Annual linear trend | 0.98 (0.78-1.23) | 1.11 (0.96-1.27) | 1.09 (0.93-1.28) | 1.06 (0.9-1.25) |

\*p <0.05; \*\*\*p <0.001
PHC, primary health center; HSS, health system strengthening; RoD, rest of the district.

## Discussion

In rural settings of sub-Saharan Africa, geographic barriers are among the main factors preventing the use of maternal health services and contributing to high rates of maternal mortality. While the effect of financial and health system barriers to maternal care can be more directly addressed via supply-side interventions, a better understanding of geographic access to care for pregnant women is necessary to devise appropriate interventions and policies. Here, we analyzed the spatio-temporal changes in geographical accessibility to health facilities for antenatal, delivery and postnatal care services in a rural district of Madagascar with substantial geographic barriers that has benefited from recent support to improve financial access and health system readiness. Using both health system information and data from a district-representative cohort, our study revealed that maternal care access decreased exponentially with increasing distance to health centers. The general trend showed improvements due to HSS across the geographic gradient. But the only statistically significant result in this setting were observed among women living within two hours from health centers. This suggests that geographically targeted interventions are necessary to ensure access to maternal care for remote populations.

In our setting, travel time to health facilities was the strongest determinant of maternal care utilization. While the negative effect of geographic barriers on maternal care utilization has been abundantly described in Africa Sub-Saharan [2,31–33], relatively few studies have assessed their impact in the context of HSS and UHC interventions [16,17]. Despite HSS efforts in place, the limited accessibility of healthcare facilities in Ifanadiana (each health center covers an average area of 200 km^2^ and approximately 10,000 people), combined with a rugged terrain and poor transportation infrastructure meant that nearly half of pregnant women lived over 2 hours from a health facility. This group had the lowest rates of antenatal, delivery and postnatal care, ranging between 2 to 5 times less than women living within 1 hour (Tables 2-3). This could explain why maternal care access in our study area was lower in 2018 than the national average for Madagascar [23]. Sufficient geographic coverage was only observed for the first antenatal visit. As a result, emergency obstetric cases in remote areas of the district would be unable to receive timely and life-saving interventions, exacerbating the risks associated with complications before, during and after childbirth.

The use of two independent longitudinal data sources allowed us to gain a detailed understanding of local spatio-temporal changes in maternal care access and to assess the consistency between results obtained at the health center level versus those at the household level. The majority of studies on geographic barriers to maternal care use data from cross-sectional surveys [14,16,34,35]. Qualitative and community-based approaches can also provide a more in-depth understanding of women’s perceptions and barriers through a combination of semi-structured interviews and focus group discussions [36–38] However, these tend to be done at one point in time and rely on self-reporting reports. Here, we demonstrate that geographic barriers were relatively stable over time during a three-year period in both sets of analyses. Moreover, we were able to assess whether health system support had a tangible impact on different populations over time, showing that inequalities in access between the closest and furthest populations (within 1h vs. over 2h from a PHC) slightly increased for antenatal care in the HSS catchment while they decreased for delivery and postnatal care (Figure 3). With few exceptions, spatial and temporal results from these two data sources were very consistent and helped identify underserved populations in the district.

This study linked individual women from maternal consultation registers and surveys to their village of residence, and we accurately estimated their travel time to the nearest PHC through a combination of participatory mapping and fieldwork. These methods are in line with recent approaches for service area analysis in sub-Saharan Africa, which aim to analyze patients’ addresses from health facility data to help delineate their catchment areas and identify populations who may experience challenges in accessing maternal care [39,40]. Moreover, the increasing availability of detailed geographic data is allowing the integration of more realistic measures of travel time to health facilities into assessments of geographic accessibility [28,41,42] Other studies on maternal care access use composite indices that combine into a single measure either multiple types of maternal health care (antenatal, delivery and postnatal care), or multiple factors related to geographic accessibility (e.g. travel time, transportation options, and topographical constraints) [43–45]. In contrast, we analyzed each maternal care indicator separately, revealing how geographic accessibility can impact them differently.

This study presented some limitations. First, population size values obtained from the MoPH used to estimate per capita utilization were inaccurate. These data are projections made each year from national censuses conducted far apart and the expected number of pregnant women is a fixed proportion of total population size, which does not take into account temporal variations in fertility preferences or other factors. This explains why per capita rates for ANC1 were higher than 1 for many fokontany in health system analyses. Second, survey data from our population-representative cohort used information from births occurring in the previous two years, and as such did not completely align temporally with health system data. This would explain slight differences observed in the temporal trends for certain indicators in each set of analyses (Figure 3). Despite this, overall rates of utilization as well as spatial and geographic trends were very consistent between the two datasets. Third, we aimed to obtain health system data from all health centers in the district, but physical registers for one health center in the initial HSS catchment were missing, which obliged us to remove eight fokontany from our health system analysis.

## Conclusion

This study shows how a combination of geo-located health system information and population-representative data can help assess the impact of geographical barriers to maternal care in rural areas of sub-Saharan Africa. It highlights that women who live more than 2 hours from a PHC have virtually no access to maternal health services at the primary care level despite efforts in place to reduce financial barriers to care and strengthen the public health system. Future research should assess whether targeted interventions for pregnant women in remote populations can achieve significant reductions in geographic barriers to care and lead to increased use of maternal health services.

## Supplementary Information

**Additional file 1**: Spatial variation in maternal care indicators (ANC4 and postanal visits), from health system data.

**Additional file 2:** Spatial variation in maternal care indicators (ANC4 and postanal visits), from cohort data.

## Data Availability

Data are available on OpenStreetMap (https://www.openstreetmap.org) and on the Shinny app (https://research.pivot-dashboard.org/)

## Acknowledgments

They are grateful to thank Pivot Research data capture (Nadia, Cathy and Dera) and the Ministry of Public Health for the support of this study and to their commitment to the health of the population. The authors also thank the Institut National de la Statistique (INSTAT) field teams for their involvement in the district-wide population survey.

## Author’s contributions

FAI, MB, AG conceived and designed the study. FAI, MR, AG analyzed the data. FAI, AG contributed to the interpretation of the data. FAI, MR, AM, RJLR, MRF, JAK, MR, HR, BR, VR, MB, KEF, AG drafted the manuscript. All authors read and approved the final manuscript.

## Declarations

### Ethics approval and consent to participate

We obtained informed consent from all participants consented to the collection of their data. This study was approved by Madagascar National Ethics Committee (062-MSANP/CE and 041-MSANP/CERBM) and The Harvard Medical School Institutional Review Board.

### Consent for publication

Not applicable

