## Supplemental figure for "Persistence of geographic barriers to maternal care services following a health system strengthening initiative in rural Madagascar"

**Additional file 1**: **Spatial variation in maternal care indicators (ANC4 and postanal visits), from health system data.** Annualized per capita utilization out of expected pregnant women, estimated as the fokontany average from data of all health centers. A) ANC4, and B) Postnatanal care.

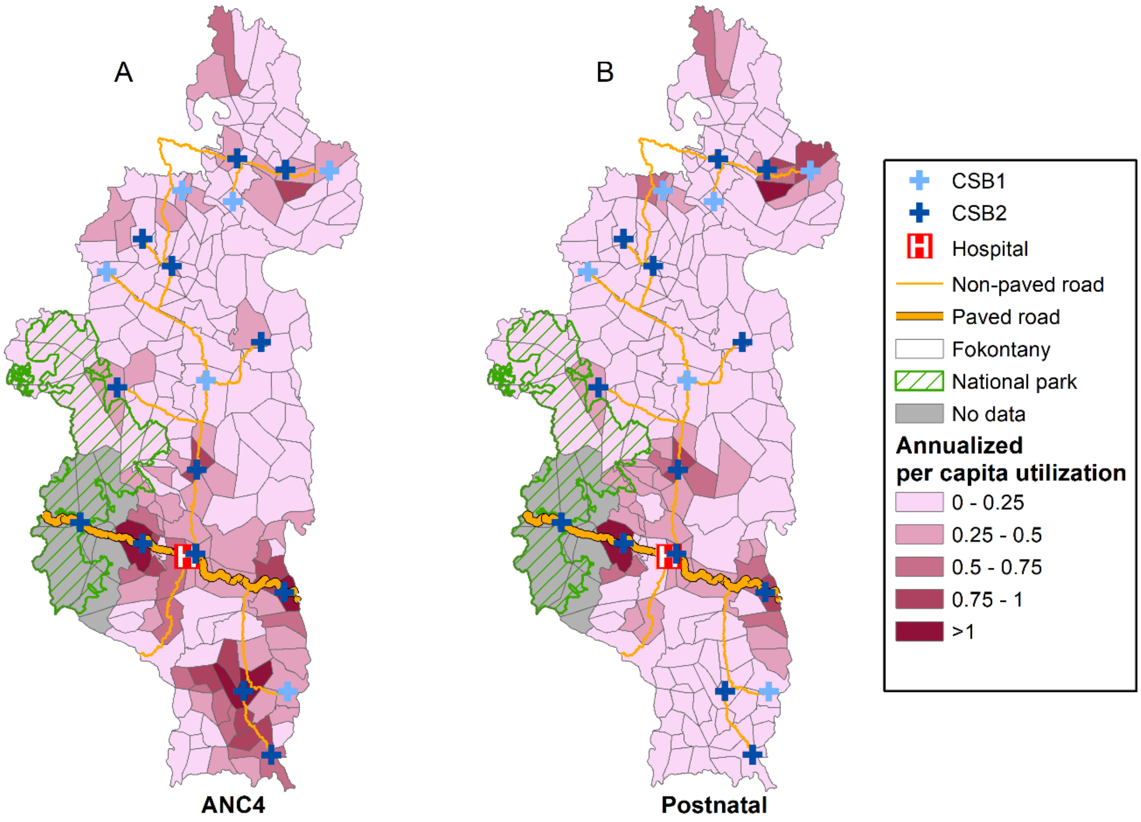

**Additional file 2: Spatial variation in maternal care indicators (ANC4 and postanal visits), from cohort data.** Percentage of pregnant women included in each of 80 clusters in cohort surveys (black circle) who accessed each maternal care service, with values interpolated using inverse distance weighted interpolation. A) ANC4, and B) Postnatal care.

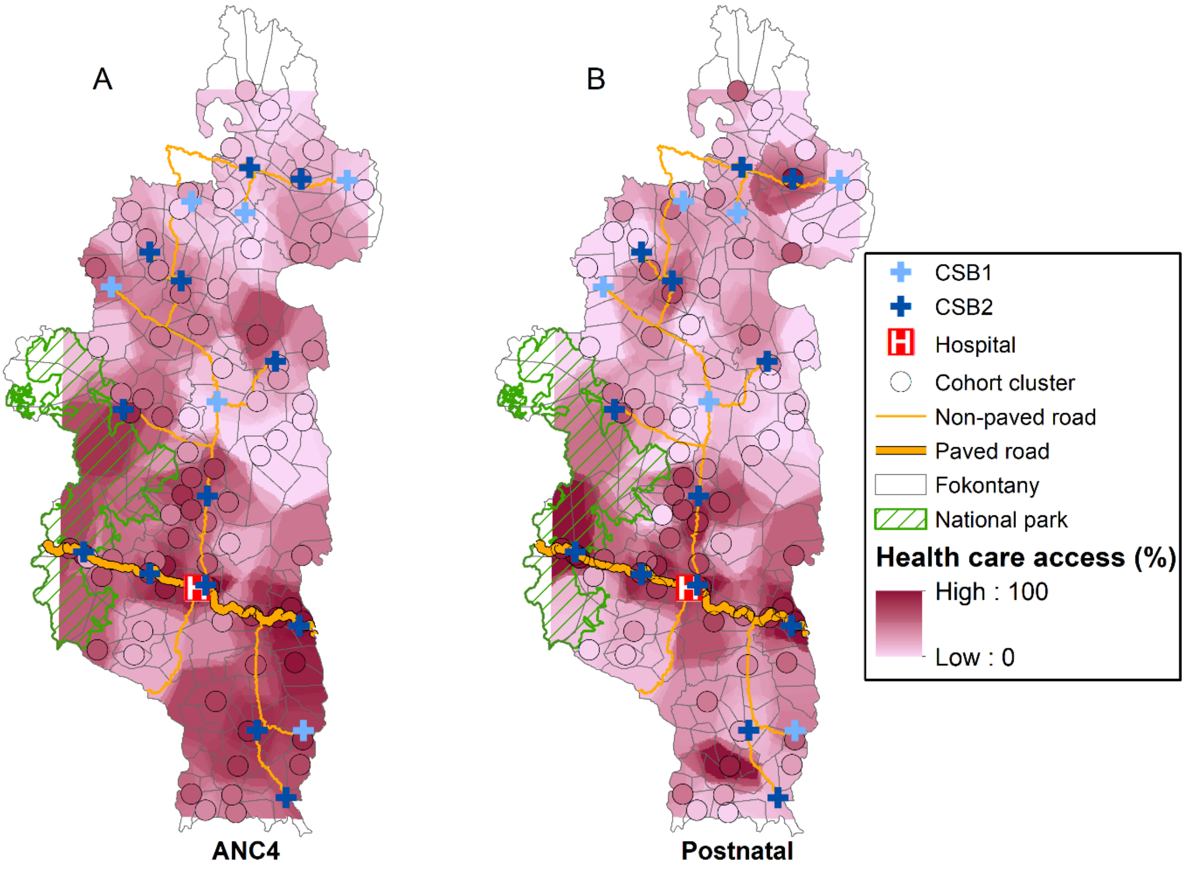
